# Laboratory Findings of COVID-19 Infection are Conflicting in Different Age Groups and Pregnant Women: A Literature Review

**DOI:** 10.1101/2020.04.24.20078568

**Authors:** Sina Vakili, Amir Savardashtaki, Sheida Jamalnia, Reza Tabrizi, Mohammad Hadi Nematollahi, Morteza Jafarinia, Hamed Akbari

## Abstract

Coronavirus disease 2019 (COVID-19), a new type and rapidly spread viral pneumonia, is now producing an outbreak of pandemic proportions. The clinical features and laboratory results of different age groups are different due to the general susceptibility of the disease. The laboratory findings of COVID-19 in pregnant women are also conflicting. Para-clinical investigations including laboratory tests and radiologic findings play an important role in early diagnosis and treatment monitoring of severe acute respiratory syndrome and coronavirus-2 (SARS-CoV-2). The majority of previous reports on the SARS-CoV-2 laboratory results were based on data from the general population and limited information is available based on age difference and pregnancy status. This review aimed to describe the COVID-19 laboratory findings in neonates, children, adults, elderly and pregnant women altogether for the first time. The most attracting and reliable markers of COVID-19 in patients were: normal C-reactive protein (CRP) and very different and conflicting laboratory results regardless of clinical symptoms in neonates, normal or temporary elevated CRP, conflicting WBC count results and procalcitonin elevation in children, lymphopenia and elevated lactate dehydrogenase (LDH) in adult patients, lymphopenia and elevated CRP and LDH in the elderly people and high CRP, leukocytosis and elevated neutrophil ratio in pregnant women.

## Introduction

Coronavirus disease 2019 (COVID-19) is a form of mild to severe respiratory disease caused by a virus belonging to the *Coronaviridae* family ^1,2^. According to the current statistics of the Johns Hopkins Coronavirus Resource Center, the disease has involved all over the world, with over 1362000 diagnosed cases and more than 76000 deaths until April 7, 2020 ^3^. In the initial phases of the disease, symptoms like fever, cough, and dyspnea can occur ^2,4^. Some patients rapidly develop acute respiratory distress syndrome (ARDS) and additional severe complications, which are ultimately followed by multiple organ failure ^5^, hence, timely diagnosis of patients is very essential. Although detection of viral nucleic acid using real-time reverse-transcription polymerase chain reaction (RT-PCR) remains the gold standard of diagnosis and monitoring, it is very time-consuming and has a high prevalence of false-negative results ^6,7^. Other laboratory tests, such as whole white blood cells (WBCs) count, neutrophil ratio, lymphocyte count, C-reactive protein (CRP), erythrocyte sedimentation rate (ESR), hemoglobin, platelets, procalcitonin, creatine kinase (CK), myoglobin, aspartate aminotransferase (AST), alanine aminotransferase (ALT), total bilirubin, creatinine, cardiac troponin I, D-dimer, albumin, lactate dehydrogenase (LDH) and several other laboratory tests have been reported to change in COVID-19 patients ^1,8-10^. The majority of previous reports on the SARS-CoV-2 laboratory results were based on data from the general population and limited information is available based on age difference and pregnancy status ^11-14^. Since the laboratory medicine provides an essential contribution to the clinical decision making in many infectious diseases including COVID-19, we reviewed a broad spectrum of COVID-19 clinical and laboratory reports to provide a comprehensive data collection in order to clarify the dark parts of laboratory-based diagnosis and monitoring of COVID-19 in a different group of patients including neonates, children, adults, elderly and pregnant women altogether for the first time.

### Search strategy

The search was performed in PubMed, Scopus and Web of Science, using the keywords “COVID-19” or “2019-nCoV” or “2019 novel coronavirus” or “SARS-CoV-2” without language or date restrictions. The title and abstract of all articles identified and those describing laboratory findings in COVID-19 patients were finally selected. The articles that had not reported data on laboratory test results were excluded. Because the aim of the study was to investigate the laboratory results among different age groups including neonates, children, adults, elderly and also pregnant women, the articles without exact clarification of age range were also excluded. Ages under 1 year, 1-17 years, 18-64 years and 65 years or older were considered as neonate, children, adults and elderly, respectively.

### Laboratory findings of COVID-19 in neonates

Clinical features of COVID-19 in neonates may be non-specific ^15^. Neonates with COVID-19 may also develop multiple organ damage and rapid illness alterations ^16^. Although laboratory data on severe acute respiratory syndrome and coronavirus-2 (SARS-CoV-2) infected neonates are still limited, there are several case reports that provide some laboratory findings. In a 6-month-old male neonate with confirmed COVID-19 infection, with no clinical signs or symptoms and high viral load, WBC count and biochemical markers including liver function tests were normal on day 2 and only neutropenia occurred on day 8 of admission ^17^. This asymptomatic neonate with normal laboratory results may play significant roles in the transmission of COVID-19 in the community. Another infected infant with no significant signs of the disease revealed different results. Blood tests showed lymphopenia, elevated liver function tests and elevated CK level ^18^. A 55-day-old female infant with only rhinorrhoea and a dry cough without fever or other significant symptoms was confirmed to have COVID-19. Laboratory tests results indicated that there were changes in hepatic function tests. Lymphocyte count, platelet count, liver function tests, procalcitonin, and serum IgM level were all slightly elevated. Other laboratory examinations including CRP, ESR, coagulation tests, hemoglobin, D-dimer and renal function tests showed no abnormalities ^16^. Laboratory examination of a 3-month-old female infected with COVID-19, suffering from diarrhea and fever with normal chest computerized tomography (CT) scan results, revealed high neutrophil levels and reduced lymphocytes ^19^. Although all the described cases were asymptomatic or with mild clinical manifestations, their laboratory results were very different and conflicting. For example, lymphocyte counts varied from lymphopenia to lymphocytosis and CRP was normal among neonates (Table 1). These findings highlight the difficulties in detecting the disease and making decisions in neonates based on symptoms and laboratory results.

**Table 1:**
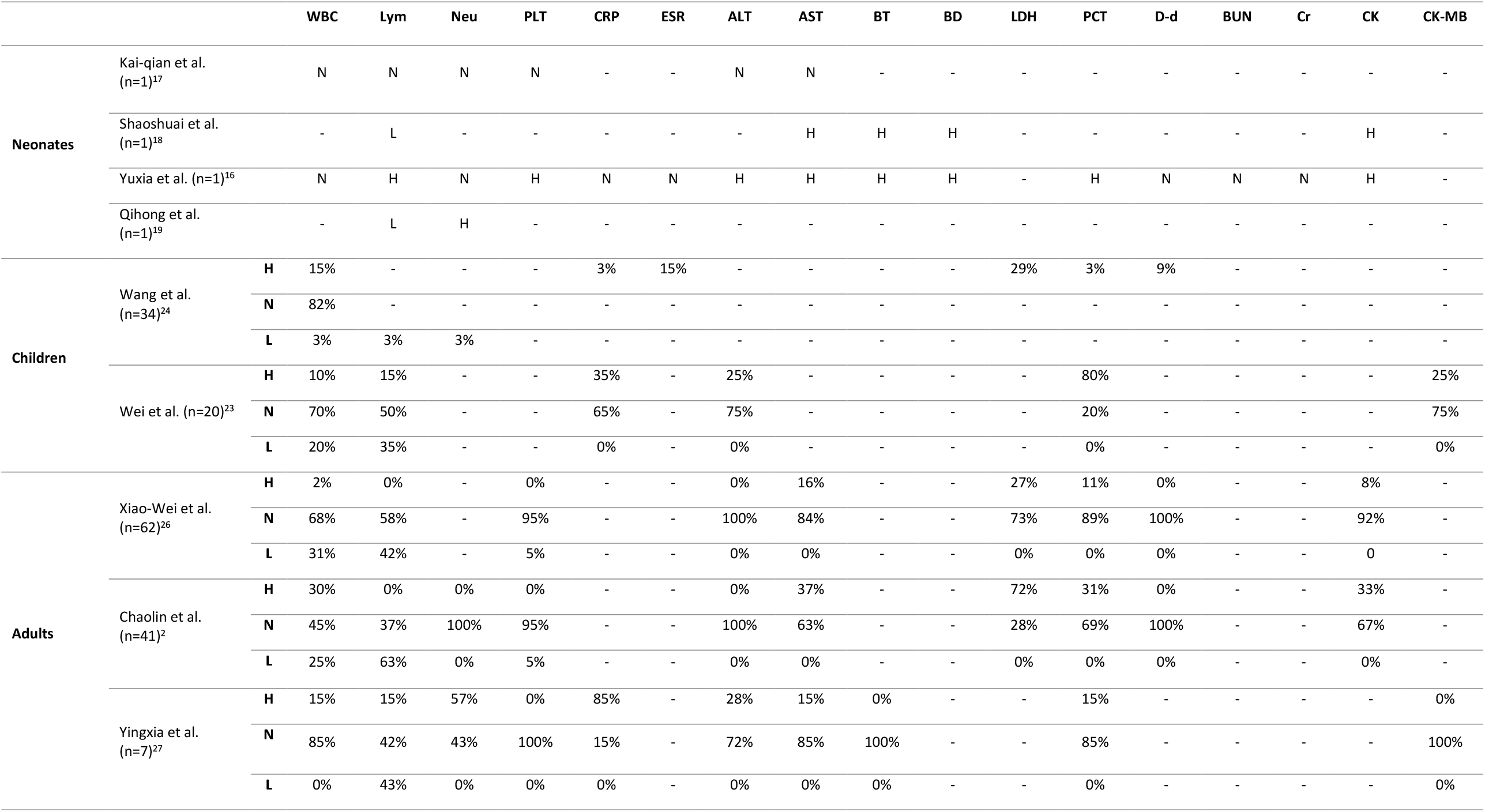
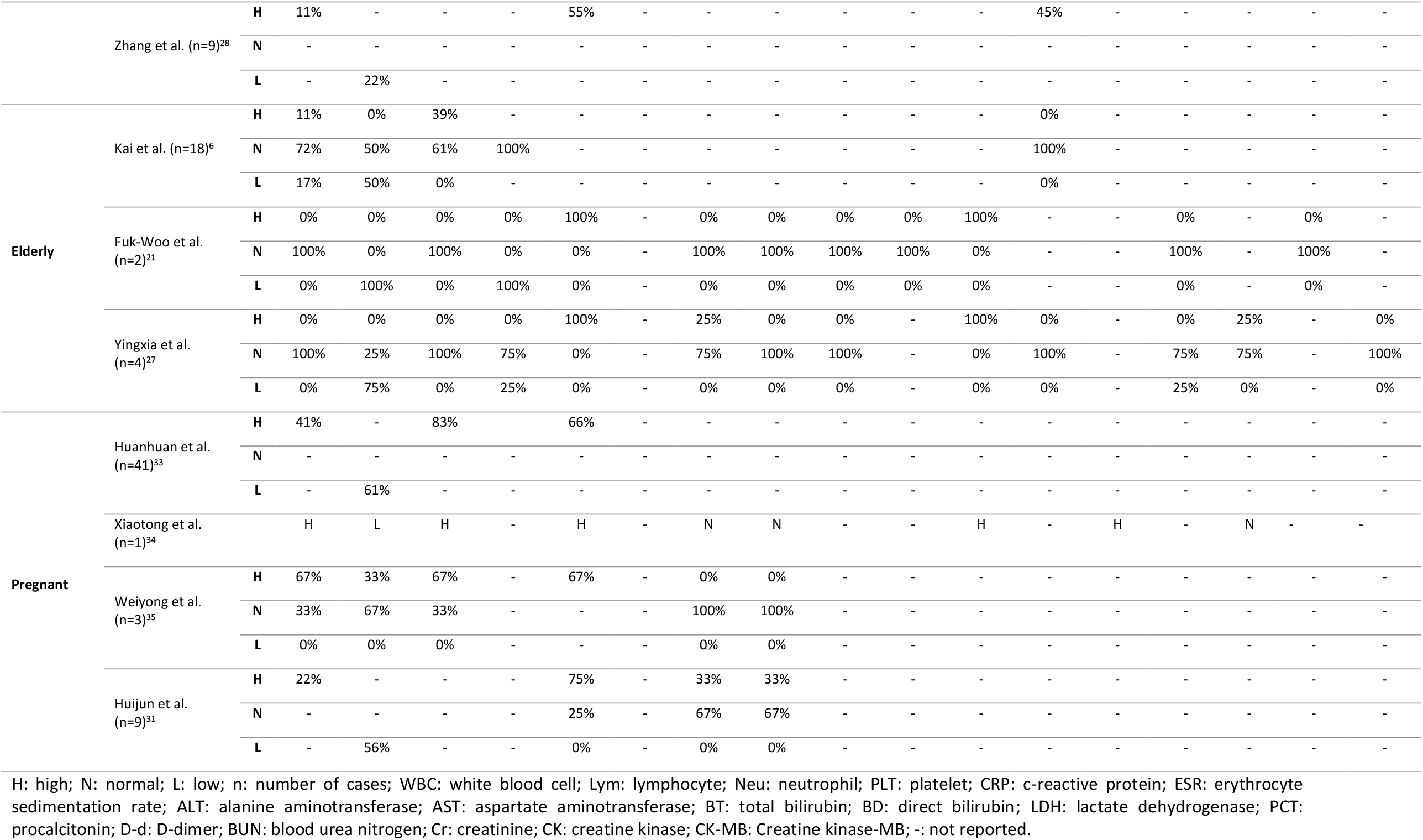
Main laboratory characteristics of the included studiess.

### Laboratory findings of COVID-19 in children

Based on current data, the majority of the SARS-CoV-2 infected children have mild or no symptoms ^20,21^. CRP was normal in the majority of infected children. WBC count results are conflicting but were normal in most of the cases ^22-25^. In a study on the 34 infected children, the WBC counts of 82% were normal. CRP levels and ESR were increased in only 1 and 5 cases, respectively. Lymphopenia and neutropenia were found in only one case, respectively. Procalcitonin elevation was found in only 1 case and D-dimer in 3 cases. Another study carried out on 10 children cases, the LDH levels were high ^24^. Wei *et al*. claimed that procalcitonin elevation was common in pediatric patients which is different from adults. The procalcitonin was elevated in 80% cases in this study which is not common in adult patients ^23^. Procalcitonin is a marker for bacterial infection and can be induced by bacteriotoxins but suppressed by interferons. It may suggest that routine antibacterial treatments may be effective in pediatric patients ^23^. Based on Wei *et al*. report and since WBC count results are very conflicting and CRP is usually normal in children, procalcitonin may be a good diagnostic marker to detect COVID-19 in children. Nevertheless, procalcitonin was reported to be elevated in only 3% of all children cases reported by Wang *et al*. ^24^. While laboratory data of infected children is limited and less than adults, normal or temporary elevated CRP, conflicting WBC count results and procalcitonin elevation are the most important laboratory reports of COVID-19 infected children (Table 1). These different laboratory results may be due to more active innate immune response, healthier respiratory tracts, and fewer underlying disorders in children as well as they are less likely to be exposed with infected people or infected environments ^22^.

### Laboratory findings of COVID-19 in adults

There are many reports regarding laboratory results of adult patients with COVID-19. However, a large number of them did not categorize the patients’ age, therefore, they were excluded from our study. In the study conducted by Xiao-Wei *et al*. on 62 COVID-19 adult patients, the laboratory results were as follows: leucopenia (31%), lymphopenia (42%), high level of LDH (27%), normal serum levels of procalcitonin (89%), normal platelet count (95%), mean levels of AST, ALT, D-dimer, and hemoglobin were within the normal range as well ^26^. In addition, Huang *et al*. reported that 63%, 25%, and 5% of patients had lymphopenia, leucopenia, and thrombocytopenia, respectively. Normal procalcitonin levels were found in 69% of adult patients. Also, LDH levels were high in 73% of patients. Creatinine, CK, and AST were elevated by 37%, 10% and 33% of patients, respectively. However, neutrophil count, prothrombin time (PT), partial thromboplastin time (PTT), albumin, hemoglobin, D-dimer, ALT, sodium, and potassium were normal among patients ^2^. In the report of Yingxia *et al*. CRP and LDH were high in 85% of patients. Fifteen percentage had leukocytosis and others had normal WBC count. Forty-three percentage had lymphopenia. The neutrophil ratio was elevated in 57% of patients. Platelet count was normal in all patients. Moreover, normal AST, ALT, and procalcitonin levels were observed in 85%, 72%, and 85% patients, respectively, while 43% showed low albumin levels ^27^. In the Mingqiang *et al*. study on nine COVID-19 adult patients, 33% had normal blood routine tests, while abnormal diagnostic results were as follows: high CRP (55%), lymphopenia (22%), leukocytosis (11%) and high procalcitonin levels (45%) ^28^. Although it seems that lymphopenia and elevated LDH are more common, the results are very different and conflicting in adult patients with confirmed COVID-19 infection (Table 1).

### Laboratory findings of COVID-19 in elderly patients

Elderly people with underlying diseases including hypertension, cardiovascular disease, and diabetes are more susceptible to COVID-19 ^6,29^. It may be due to changes in the elderly‘s lung anatomy and muscle atrophy leading to changes in respiratory system function. Because elderly people with COVID-19 are prone to multi-system organ dysfunction ^6^, they may present different laboratory outcomes. Early diagnosis of SARS-CoV-2 in elderly patients is absolutely essential because elderly patients especially those with underlying diseases are more susceptible to severe illness and the mortality of them is higher ^6,30^. A study carried out by Kai *et al*. demonstrated that lymphopenia in elderly COVID-19 confirmed patients was significantly higher than that in the young and middle-aged groups. Furthermore, CRP was significantly higher in elderly patients compared to young and middle-aged groups. Other laboratory results including WBC count, neutrophil ratio, platelet, hemoglobin, procalcitonin, albumin, and serum creatinine were not significantly different between the study groups ^6^. In the study of Fuk-Woo *et al*. lymphopenia, low platelet count and elevated CRP were observed in all elderly patients. LDH, activated PTT and fibrinogen were also elevated in all elderly patients. Similar to Kai *et al*. results, WBC count, neutrophil ratio, hemoglobin, albumin, and creatinine were normal. Liver function tests, CK, urea, and PT were also within the normal range ^21^. In Liu *et al*. report, CRP and LDH were high in all elderly patients. Lymphopenia also detected in most of the old cases. WBC count, albumin, liver enzymes, myoglobin, CK-MB, procalcitonin, blood urea nitrogen (BUN), creatinine, and platelets were almost normal in all cases ^27^. Taken together, it is safe to claim that lymphopenia and elevated CRP and LDH are tests that should be considered more particularly in elderly COVID-19 people (Table 1).

### Laboratory findings of COVID-19 in pregnant women

Pregnant women are more susceptible to SARS-CoV-2 and severe pneumonia, because they are at physiological adaptive changes and immunosuppressive state during pregnancy ^31,32^. An ample body of evidence has revealed that the clinical symptoms and laboratory findings of infected pregnant women are atypical in comparison with the non-pregnant adults. The leukocytosis and elevated neutrophil ratio were reported to more common in the COVID-19 infected pregnant women in the Liu *et al* study ^33^. However, there was no significant difference regarding lymphopenia among pregnant and non-pregnant groups. CRP elevation is obvious in most of the cases ^33^. In the study of Wang *et al*., laboratory findings included high leukocyte count, elevated neutrophil ratio, lymphopenia, and elevated CRP, D-dimer, and LDH. However, aminotransferase levels and creatinine were reported in a normal range ^34^. Moreover, LIU *et al*. reported that leukocytosis and elevated neutrophil ratio, elevated CRP and interleukin 6, and low albumin are the main laboratory findings of pregnant women, while the normal amount of ALT, AST, ferritin, and ESR were reported ^35^. Chen *et al*. reported that among nine pregnant women with COVID-19 pneumonia, 55% had lymphopenia. 66% had elevated concentrations of CRP and 33% had increased concentrations of ALT and AST. Additionally, inconsistent with other reports indicated leukocytosis and elevated neutrophil ratio, 77% of patients had a normal WBC count ^31^. It seems that high CRP, leukocytosis and elevated neutrophil ratio are the most reliable markers of COVID-19 among pregnant women and other laboratory tests are conflicting (Table 1). Nevertheless, the physiological findings regarding leukocytosis and elevated neutrophil ratio due to adaptations to gestation ^33^ can complicate the situation. Comprehensive data on larger population of pregnant women with COVID-19 are needed to better understanding the impact of SARS-CoV-2 on maternal and birth outcomes.

## Conclusion

There is a crucial need to better recognize the full laboratory spectrum of COVID-19 in the different populations in order to help early diagnosis of the disease. This review aimed to describe the COVID-19 laboratory findings in neonates, children, adults, elderly and pregnant women altogether for the first time in order to better comparison of laboratory results and provide an insight about the diagnosis of the disease in different age populations. We showed that the laboratory findings of COVID-19 confirmed pneumonia patients were very different and conflicting, however, some tests are attracting more attention. Laboratory results of neonates were very different and conflicting regardless of clinical symptoms, and CRP was normal in the cases compared to other age categories though. Normal or temporary elevated CRP, conflicting WBC count results and procalcitonin elevation are the most important laboratory reports of COVID-19 infected children. With respect to adult patients, lymphopenia and elevated LDH were more common. Lymphopenia and elevated CRP and LDH were markers that should be considered more in elderly people. High CRP, leukocytosis and elevated neutrophil ratio were the most reliable markers of COVID-19 among pregnant women. Differences in the distribution, functioning and maturation of viral receptors is often mentioned as a probable reason of the age-related differences in clinical and laboratory features on COVID-19 patients ^15^. There were many limitations including lack of age range clarification in many reports, the low number of studies in neonates, children and pregnant women, small sample size of some studies, different or lack of normal ranges of laboratory tests and incomplete laboratory data reporting in previous studies.

## Data Availability

The dataset of the current study are available from the corresponding author on reasonable request.

## Abbreviations

WBC: white blood cell
COVID-19: coronavirus disease 2019
ESR: erythrocyte sedimentation rate
SARS-CoV-2: severe acute respiratory syndrome coronavirus-2
CK: creatine kinase
AST: aspartate aminotransferase
ALT: alanine aminotransferase
LDH: lactate dehydrogenase
RT-PCR: real-time reverse transcription-polymerase chain reaction
ARDS: acute respiratory distress syndrome
CRP: C-reactive protein
PT: prothrombin time
PTT: partial thromboplastin time
BUN: blood urea nitrogen

## Declarations

## Acknowledgements

Not applicable.

## Authors’ contributions

All authors contributed in collecting data and writing the manuscript. All authors have read and approved the manuscript

## Availability of data and materials

The dataset of the current study are available from the corresponding author on reasonable request.

## Ethics approval and consent to participate

Not applicable.

## Consent for publication

Not applicable.

